# The impact of an ambient documentation tool on the care experience of clinical pharmacists embedded in ambulatory clinics: A mixed methods evaluation

**DOI:** 10.64898/2025.12.19.25342691

**Authors:** Anthony W. Olson, Apoorva Pradhan, Payton Whary, Sarah Krahe Dombrowski, Duncan Dobbins, Theron Ward, Kimberly Obert, Cory N. Siegrist, Eric A. Wright

**Affiliations:** Geisinger – College of Health Sciences, Department of Pharmacy, Danville, PA, USA; Geisinger – College of Health Sciences, Department of Investigational Medicine, Danville, PA, USA; Geisinger - Center for Pharmacy Innovation and Outcomes, Danville, PA, USA; Geisinger - Center for Kidney Health Research, Danville, PA, USA; Geisinger – Enterprise Pharmacy, Danville, PA, USA; Geisinger – Digital Transformation Office, Danville, PA, USA

**Author notes:** Corresponding Author: Anthony W. Olson, PhD, PharmD, 100 N Academy Ave, Danville, PA 17822.

**Keywords:** ambient documentation, artificial intelligence, documentation burden, burnout, care experience, clinical pharmacists

## Abstract

**Purpose:** Ambient Documentation Tools (ADT) are an emerging technology designed to help clinicians complete documentation better with less time and effort. This study aimed to understand the impact of ADT on the pharmacist care experience.

**Methods:** Data from Epic Signal, surveys, and interviews were collected between February 2024 and October 2025 from 41 medication therapy disease management (MTDM) pharmacists across 33 primary care and subspecialty clinics at a large integrated health system who were given ADT licenses. Study variables included pharmacist ADT utilization rate and pre-/post-changes in time spent in notes per encounter, as well as perceptions of documentation burden, patient access, undivided attention for patients, afterhours documentation, and burnout. Quantitative data were analyzed using descriptive statistics, regression models, and comparative tests of pre-/post-statistical significance and effect size. Qualitative data were mined for exemplary excerpts to deepen understanding.

**Results:** Thirty pharmacists from 28 clinics utilized ADT and provided usable responses. ADT was utilized for 65% of eligible encounters, and the average time in notes per encounter fell 86 seconds post-ADT. Pharmacist perceptions of documentation burden, undivided attention ability, and afterhours documentation improved post-ADT. Interview responses were largely positive for most variables and revealed multiple explanatory mechanisms.

**Conclusion:** ADT meaningfully improved several care experience aspects for MTDM pharmacists over a short period of time (2-7 months). Future research with larger samples and longer time horizons across multiple health systems is needed to investigate the full and sustained impact of ADT on the care experience.

**Key points:**

- This mixed-methods study is the first to investigate the impact of an ambient documentation tool (ADT) on the care experience of clinical pharmacists embedded in ambulatory clinics.
- Data from Epic Signal, surveys, and interviews were triangulated to assess ADT usage and evaluate pre-/post-ADT deployment changes in pharmacist care activities and perceptions.
- Pharmacists activated ADT for 65% of eligible encounters, reducing time spent in notes by 86 seconds per encounter and improving their perceptions of documentation burden, ability to give patients undivided attention, and afterhours documentation.

## INTRODUCTION

Clinical pharmacists in ambulatory settings devote a sizable share of each workday to documentation activities in the electronic health record (EHR) for their clinical encounters, often extending after regular clinic hours.^1,2^ The amount of time and effort (T&E) that clinicians spend working in the EHR has been associated with burnout and lower quality for some aspects of care.^3,4^ Additionally, clerical tasks like documentation can occupy cognitive bandwidth that reduces working capacity for tasks like clinical reasoning and communicating with patients.^5^

Recent advancements in artificial intelligence have led to ambient documentation tools (ADT) designed to reduce clinician T&E in the EHR.^6,7^ ADT audio-records care encounters between patients and clinicians, and then auto-generate notes in the EHR for review by clinicians. Emerging evidence suggests ADT may help clinicians document faster and better (e.g., improved comprehensiveness, readability, and accurate capture of service levels for billing), decreasing screen time in the EHR, and increasing face time with the patient.^8–11^ The anticipated impact includes a better care experience for both the patient and clinician and more opportunity for high-quality, person-centered care.

Despite encouraging data in the medical literature, no studies examine the impact of ADT on pharmacist-led medication management services. Pharmacists’ workflow, practice activities, documentation, and billing models differ from other health professions and should be evaluated accordingly.^12–15^ In February 2025, Geisinger began pilot testing an ADT with pharmacists practicing medication therapy disease management (MTDM) in primary care and subspecialty ambulatory practices. The aim of this study is to understand the impact of ADT on the pharmacist care experience in medication management encounters. The objectives were to: (1) describe pharmacists’ ADT ‘Utilization Rate’ and ‘Time Spent in Note per Encounter’ pre-/post-ADT deployment; and (2) evaluate the effect of ADT on pharmacists’ perceptions of ‘Documentation Burden,’ ‘Patient Access,’ ‘Undivided Attention,’ ‘Afterhours Documentation,’ and ‘Burnout.’

## METHODS

### Study Design, Participants, and Setting

This pre-post mixed-methods study was conducted at Geisinger, a large integrated health system serving over one million patients in central and northeastern Pennsylvania. Geisinger’s MTDM program was established in 1996,^16^ and as of October 2025 employs 84 clinical pharmacists embedded in ambulatory clinics with collaborative practice agreements who conduct approximately 210,000 medication management encounters per year across 68 primary care and subspecialty sites.

On 25 September 2024, Geisinger deployed an ADT developed by Abridge^17^ to a group of outpatient physicians. On 19 February 2025, the deployment was extended to four ambulatory pharmacists conducting MTDM services. An additional six MTDM pharmacists were added on 16 April 2025, followed by another three MTDM pharmacists on 13 May 2025. The three waves totaling 13 pharmacists embedded in 10 clinics represented a purposive sample of varying primary care and subspecialty clinics to capture practice model differences. On 15 July 2025, a fourth wave of 28 pharmacists across 23 clinics representing all Geisinger MTDM pharmacists with an iOS device who requested ADT were provided Abridge licenses.

### Intervention

Abridge is an ADT with “out-of-the-box” functionality requiring minimal clinician training and Epic Haiku configuration (mobile EHR application). Abridge is compatible with iOS and Android devices, but Abridge licenses were available to Geisinger clinicians with the former during the study.

When a pharmacist receives patient consent to use Abridge, the pharmacist hits a button on their iOS device to activate an ambient recording of the interaction. Abridge’s large language model then transforms the collected voice-to-text data in real-time into a structured clinical note draft in the EHR (Supplement A). Clinicians must review, edit, and approve drafts for appropriateness and accuracy before adding notes to the patient’s medical record. Audio recordings are deleted 30 days later to protect private health information.

### Data Collection and Variables

Data were collected from three sources: Epic Signal (analytics tool tracking user EHR activities);^18^ online surveys; and semi-structured interviews. Quantitative data from Epic Signal and survey sources were collected in Waves 1-4, and qualitative data from interviews were collected in Waves 1-3. Data for participant demographics (sex, age, race/ethnicity) and practice-related characteristics (primary care vs. subspeciality, years of MTDM experience) were collected with interviews for Waves 1-3, and via online surveys using REDCap for pharmacists in Wave 4.^19^Supplement B presents operational definitions for all study variables described below.

For objective 1, Epic Signal data were obtained from the EHR for all participants between February 2024 and October 2025 to determine ADT ‘Utilization Rate’ and pre-/post- ‘Time Spent in Note per Encounter.’^18^ ‘Utilization Rate’ was reported as the percent of all ADT-eligible encounters during the participating pharmacists’ study periods for which ADT was activated. ‘Time Spent in Notes per Encounter’ was reported as the percentage pre-/post-ADT change in the average number of minutes spent in notes. Each pharmacist’s pre-/post- comparison periods were equivalent (e.g., 3 months pre and 3 months post), but inter-pharmacist comparison periods varied by participant Wave (i.e., Wave 1=7 months, Wave 2=5 months, Wave 3=4 months, Wave 4=2 months). The ‘Time Spent in Notes per Encounter’ variable was aggregated for each individual pharmacist within a defined study period (i.e., pre- or post-) that divided the minutes in Epic spent on “Note and Letter” activities by the number of ADT-eligible encounters.

For objective 2, pre-/post-ADT survey data were collected between February 2025 and September 2025 via electronic, self-administered Typeform questionnaires.^20^ Pharmacists were sent email invitations to baseline surveys pre-ADT license activation. Email invitations to the follow-up survey were sent to pharmacists 30+ days post-ADT activation.

Pre-/post- surveys contained eight items that collected data for five study variables: ‘Documentation Burden,’ ‘Patient Access,’ ‘Undivided Attention,’ ‘Afterhours Documentation,’ and ‘Burnout.’ ‘Documentation Burden’ was a composite variable from the NASA Task Load Index (NASA-TLX)^21^ that summed three distinct 21-point Likert-type items (0=minimum, 20=maximum) for pharmacists’ perceptions of their effort level, mental demand, and temporal demand. The variables of ‘Patient Access’ and ‘Undivided Attention’ were measured using 5-point Likert-type items (*1=strongly disagree, 5=strongly agree*). The ‘Afterhours Documentation’ variable measurement asked pharmacists to estimate the average time they spent per week writing notes outside clinic hours. The ‘Burnout’ variable was measured using a 5-point Mini-Z worklife and burnout reduction instrument (*1=no burnout; 5=complete burnout)*.^22^

Qualitative data were collected from interviews conducted via virtual meetings with pharmacists by AWO (male) and PW (female) between 15 July 2025 and 28 August 2025. Pilot-tested interview guides (Supplement C) explored the underlying factors, reasoning, and situations informing pharmacists’ survey responses and ADT utilization. Interviews were audio-recorded and transcribed with Microsoft Teams, and interviewers discussed their field notes and observations afterwards. Interviewers had an established working relationship with study participant and co-author SKD.

### Data Analysis

Descriptive statistics were calculated for all quantitative study variables, but data from pharmacists not completing the pre- and post- surveys were excluded. The average ‘Time Spent in Notes per Encounter’ per pharmacist was computed with Epic Signal. The unadjusted average difference between pre-/post- ‘Time Spent in Notes per Encounter’ was computed using the Wilcoxon signed rank-test. We evaluated the association between ‘Time Spent in Notes per Encounter’ and pharmacists’ characteristics, including age, gender, race, ethnicity, years of MTDM experience, and time since ADT license activation. Associations were assessed in two steps: (1) Least Absolute Shrinkage and Selection Operation (LASSO) for variable selection, then (2) a generalized estimating equations (GEE) model with identity link and clustered at the pharmacist level. The optimal model selected was a seeded LASSO model with the lowest AICC model fit parameter identified age, sex, practice setting, duration since ADT-initiation, and years of MTDM experience. A GEE model subsequently assessed associations between time spent in notes pre-/post-initiation and adjusted for the best fit LASSO model variables.

For survey data, the means for continuous variables were compared using paired t-tests for normally distributed data and paired Wilcoxon signed-rank tests for non-normally distributed data. Cohen’s d value was calculated for the survey to quantify variation of the effect size (<0.5=small, 0.5-0.8=moderate, >0.8=large). The burnout variable was converted to a binary variable (scores <3 and ≥3) and analyzed with chi-square test and logistic regression to calculate odds ratios for pre-/post-ADT periods. Analyses were performed using SAS Enterprise Guide 8.3.

Quantitative findings were triangulated with exemplar excerpts identified by AWO and PW using Atlas.ti to represent the range of factors, reasoning, and situations underlying pharmacist activities and perceptions.^23,24^ AWO and PW modified exemplar excerpts for readability as follows: deletion of repeated words/phrases or filler words (e.g., umm, ahh); grammatical corrections; spelling-out acronyms; word/phrase substitutions in parentheses to improve specificity/clarity or protect anonymity; and ellipses to bridge aligned content from different parts of the interview. Interviews reviewed all modified exemplar excerpts and confirmed preservation of their intended meaning.

### IRB Determination

The Geisinger IRB determined this study met criteria for human subjects research exemption (Study#2025-0556). Some quantitative data were collected as part of a quality improvement pilot, and reporting adhered to the Standards for Quality Improvement Reporting Excellence 2.0 guidelines.^25^

## RESULTS

Of 41 pharmacists in Waves 1-4 meeting eligibility criteria, 40 used ADT at least once during the study. Of these, 30 pharmacists from 28 clinics submitted usable survey responses (completed all study surveys that could be combined with Epic Signal data), representing a 73% response rate. Of the 13 pharmacists in Waves 1-3 invited to interview, 12 accepted and provided qualitative data for analysis (92% response rate).

**Table 1** presents the demographic profile of study participants across the quantitative (Waves 1-4) and qualitative (Waves 1-3) data sets. Waves 1-4 were predominately female (80%), Millennial (73%), and White (n=30). The average years of MTDM experience for pharmacists was 7.33 and most were in family practice clinics (59%). Waves 1-3 were predominately female (67%), Millennial (83%), and White (n=30). Pharmacists averaged 8.88 years of MTDM experience and were primarily in family practice clinics (47%).

**Table 1.** Pharmacist Demographic and Practice-related Characteristics by Data Type.

| Characteristic | Waves 1-4 (Quan data)<br>n=30 | Waves 1-3 (Qual data)<br>n=12 |
| --- | --- | --- |
| Sex at birth (n, %) |  |  |
| Female | 24, 80% | 8, 67% |
| Male | 6, 20% | 4, 33% |
| Age (n, %) |  |  |
| 21-27 (*Generation Z*) | 4, 13% | 0, 0% |
| 28-44 (*Millennial*) | 22, 73% | 10, 83% |
| 45-66 (*Generation X*) | 4, 13% | 2, 17% |
| *All ages (Average, SD)* | 35.2, 8.40 | 37.33, 8.11 |
| Race/ethnicity (n) |  |  |
| Asian | 1, 3% | 1, 8% |
| White | 30, 100% | 12, 100% |
| Clinic practice setting (FTE, %) |  |  |
| Primary Care - Family Practice | 17.65, 59% | 5.65, 47% |
| Primary Care - Geriatric | 3.4, 11% | 3.4, 28% |
| Subspecialty - Cardiology | 2, 7% | 0 |
| Subspecialty - Endocrinology | 3, 10% | 1, 8% |
| Subspecialty - Pain/Behavioral Health | 2.95, 10% | 0.95, 8% |
| Subspecialty - Rheumatology | 1, 3% | 1, 8% |
| MTDM experience (average years, SD) | 7.33, 5.09 | 8.88, 3.99 |

Note: All participants providing qualitative data also provided quantitative data and are represented in both data type groups. Three pharmacists practiced in multiple settings, thus the n for “clinical practice setting” represents FTE distribution. Race/ethnicity n exceeded N and % summed to >100% to account for participants who self-identified in more than one category. Percentages may not sum to 100 due to rounding. Abbreviations: Quan=quantitative, Qual=qualitative, SD=standard deviation, FTE=full-time equivalency, MTDM=medication therapy disease management.

**Table 2** displays ADT ‘Utilization Rate’ and ‘Time Spent in Note per Encounter.’ ADT was activated for 65% of eligible medication management encounters, with over two-thirds of pharmacists using ADT between 25% and 75% of these encounters. The best fit GEE model associated significant increase in ADT utilization for encounters with being male (p=.048), primary care settings (p=0.016), and fewer years of MTDM experience (p=0.004). Qualitative data suggested ADT was most valuable in longer visits addressing comorbidities with ADT-drafted notes embedded in SmartText templates. Reasons for non-use of ADT included shorter visits, using SmartPhrases with discrete options, learner-conducted encounters, anticipation of patient refusal, incompatibility with some telemedicine encounters, or forgetting to bring their iOS phone into in-person encounters.

**Table 2.** Pharmacist Utilization and EHR Time (N=30)

| Variable (unit) | Pre-ADT | Post-ADT | p | Effect size | 95% CI |
| --- | --- | --- | --- | --- | --- |
|  | average (standard deviation) |  | Wilcoxon rank sum | GEE estimate |  |
| *Exemplar Excerpt* |  |  |  |  |  |
| Utilization rate (%) | -- | 64.8 (21.8) | - | - | - |
| • “I have [an Abridge] license on my phone. I don't feel comfortable leaving my personal device with a student ...[Also] I would want the student to be able to work on their documentation skills [without Abridge] as well. You know, I don't want them to think that wherever they go in practice that they wouldn't have to document...If I am working from home and am completing a scheduled telephonic medicine visit for patients unable to complete a video or in-person visit, I also am not able to do Abridge from the same device.” -Millennial, 6yrs experience, subspecialty practice<br>• “Some [patients] don't want [Abridge to be used]. I have had few and far between that don't allow it, but there's occasionally somebody that doesn't want to have it. So, you know we don't use it. I think the other thing [is] I don't have [SmartText] templates for all of my notes, so sometimes it's just not feasible to use [Abridge].” -Generation X, 8.5yrs experience, primary care practice<br>• “If I [don't] remember to bring my phone in with me to the exam room, that is really the only reason why [I don't use Abridge] ...I mean, I haven't had any patients decline, you know, using Abridge.” -Millennial, 11 yrs experience, primary care practice |  |  |  |  |  |
| Time Spent in Notes per Encounter<sup>a</sup> (minutes) | 14.32 (5.45) | 12.89 (6.11) | <0.001 | -1.43 | -2.23 to -0.63 |
| • “Interesting. It's less time savings than I would have thought. I'd be curious what the time means. Is it the time I had a chart open, for example?...I would have said it was saving me significant time, um, in note writing and improving the quality both of the notes.” -Millennial, 11yrs experience, primary care practice<br>• “It's amazing how I'm able to [make] corrections and in a matter of like 2-3 minutes flat, I'm able to close that chart. So something that used to take maybe 15 minutes to manually write the note, now it's taking like 2 to 3 minutes and everything is written out perfectly and documented well.” -Millennial, 11yrs experience, primary care practice<br>• “I can do a lot in a few minutes. I can check my emails, I can talk to a doctor about another patient, I can check an impact message and send a [patient portal] message. So I feel like on the surface it may not seem like a lot [of time savings from Abridge], but also [it adds up] when you have at least 10 visits a day.” -Millennial, 5yrs experience, primary care and subspecialty practice |  |  |  |  |  |

<sup>a</sup>The average change in minutes spent in note per encounter are obtained from an adjusted regression model for age, the number years practicing as an MTDM pharmacist, and time since initiation of ambient usage. Demographic data were not available on one pharmacist, who was thus excluded from the regression model. Abbreviation: ADT=Ambient documentation tool, GEE=Generalized Estimating Equations model, yrs=years

The adjusted model showed that the average ‘Time Spent in Notes per Encounter’ for ADT-eligible encounter types decreased by 86 seconds (-9.9%) between the pre-/post-intervention study periods (GEE -1.43, 95%CI -2.23 to -0.63, p<0.001). Interview data suggested most pharmacists felt the 86 seconds of time-savings per encounter was underestimated.

**Table 3** presents quantitative pre-/post-ADT comparisons for five survey variables accompanied by qualitative excerpts that exemplify the rationale underlying responses. Results were statistically significant for all variables except Patient Access (p=0.6621).

**Table 3.**
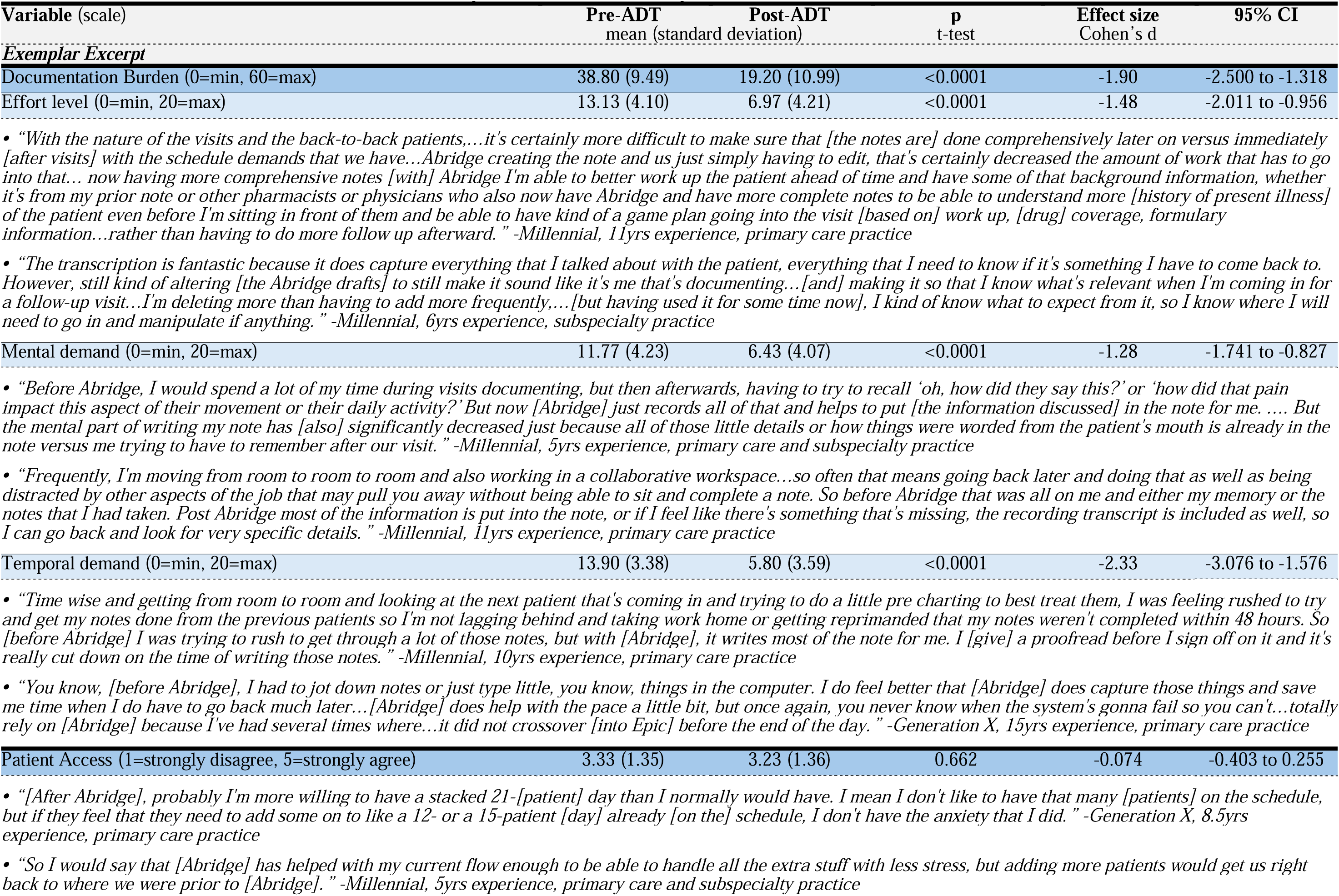
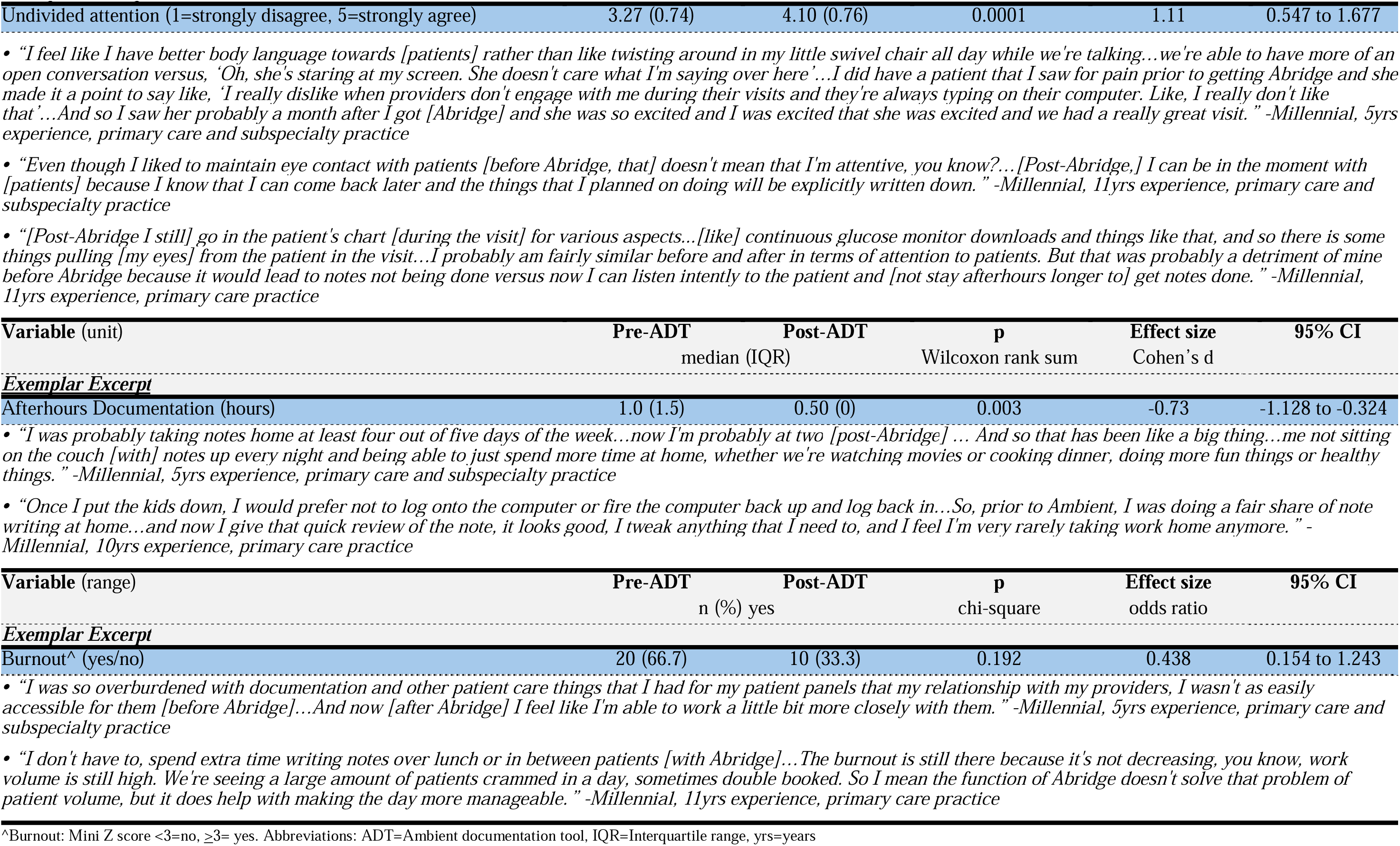
The Effect of ADT on Pharmacist Perceptions of the Care Experience (N=30)

On average, pharmacist pre-/post-ADT perceptions of ‘Documentation Burden’ were halved (mean Δ: -50%, Cohen’s d: -1.90, 95%CI: -2.5 to -1.318) with large effect sizes across its component variables of effort level (mean Δ: -47%, Cohen’s d: -1.48, 95%CI: -2.011 to -0.956), mental demand (mean Δ: -45%, Cohen’s d: -1.28, 95%CI: -1.741 to -0.827), and temporal demand (mean Δ: -58%, Cohen’s d: -2.33, 95%CI: -3.076 to -1.576). Most pharmacists interviewed found proofreading and editing ADT-drafted notes easier than writing a note. However, a few pharmacists expressed concern about errors in ADT-drafted notes (e.g., typos, inaccuracies) they wouldn’t have made themselves. Some pharmacists attributed the lower levels of effort and mental demand when documenting to ADT resolving their difficulties recalling what they or the patient said during the encounter, especially amidst juggling other responsibilities or back-to-back patients preventing note completion. Several pharmacists shared ADT-drafted notes were more comprehensive than their self-written notes and especially reduced T&E searching the EHR on follow-up for patients with complex comorbidities. Most pharmacists also expressed feeling less rushed to complete their notes except for pharmacists who self-described as fast-typers or strong multitaskers.

The average perceived capacity to provide ‘Patient Access’ if urgently needed slightly decreased post-ADT (mean Δ: -3%, Cohen’s d: -0.074, 95%CI: -0.403 to 0.255) but was not statistically significant (p=0.662). While no pharmacists reported that ADT interfered with their patient access, very few reported they could add another patient to their schedule if urgently needed. One pharmacist stated that increasing their patient panel size would negate their perception of the intervention’s benefit.

On average, ‘Undivided Attention’ improved post-ADT at a large effect size (mean Δ: +25%, Cohen’s d: 1.11, 95%CI: 0.547 to 1.677). Many interviewed pharmacists reported that ADT facilitated more patient eye-contact in encounters and less time staring at screens. One pharmacist reporting pre-/post-ADT self-ratings of 4 (agree), shared that their high level of undivided attention remained unchanged, but no longer at a time cost. Other self-described fast-typers or strong multi-taskers also showed little-to-no change and reported time spent in the EHR was necessary for high quality care and didn’t affect their attention level. Several pharmacists reported using the EHR differently post-ADT, such as searching for information to address care gaps instead of paraphrasing patient quotes or writing self-reminders.

The number of ‘Afterhours Documentation’ per week reported by pharmacists reduced from 1 hour to 0.5hrs, representing a moderate effect size (Cohen’s d: -0.73, 95%CI: -1.128 to - 0.324). Several pharmacists expressed in interviews how ADT meaningfully reduced or eliminated their afterhours documentation that otherwise would have led to delinquent note completion or less quality time with family. However, pharmacists who described themselves as efficient documenters did not report this benefit.

The odds of burnout were statistically insignificant pre-/post-ADT (odds ratio: 0.438, 95% CI: 0.154 to 1.243, p=0.1208). Most qualitative excerpts reaffirmed little-to-no burnout changes post-ADT deployment but credited ADT for having less stress, more manageable days, fewer working lunches, or opening time for collaboration with colleagues.

## DISCUSSION

This mixed-methods study is the first to evaluate the impact of a turnkey ADT on the pharmacist care experience in medication management encounters. Findings show pharmacists spent less time in notes per encounter post-ADT, corresponding with improved perceptions of ‘Documentation Burden,’ ability to give patients ‘Undivided Attention,’ and less ‘Afterhours Documentation.’ The results suggest that ADTs with performance capabilities approximating Abridge can meaningfully reduce several care experience burdens relatively quickly (2-7 months).

### Pharmacist Utilization and Time Spent in EHR

The 65% ‘Utilization Rate’ within 2-7 months is slightly above the range of existing studies of ADT implementation in medicine, which range between 20 to 55%.^8,9,26,27^ The finding of increased of ADT utilization in encounters with being male and fewer years of MTDM experience were not comparable with the medical literature, which lacked studies reporting differences in ADT-utilization by gender. However, some physician studies suggested female physicians spend more time documenting and others were composed many long-practicing clinicians adopting ADT.^8,28,29^ More investigation is needed to reconcile the ambivalent findings regarding clinician sex and practice experience with ADT-utilization given the selection bias limitations of our study design. The increased ADT-utilization in primary care pharmacist encounters aligned with the medical literature, which attributed the finding to primary care having fewer specialty-specific needs.^8,10,30^ Qualitative findings also raised two important literature gaps for best practices for ADT-utilization: student/resident training and patient acceptance. First, ADT-usage by trainees risks bypassing learner skill development in critical thinking, reflection, and patient communication.^31,32^ Second, patient willingness for ADT-use may depend on how the technology is described at consent. One study found consent rates dropped from 82% to 55% when patients received detailed information about ADT capabilities and data privacy rules.^33^

The average ‘Time Spent in Notes per Encounter’ post-ADT decreased by 86 seconds, aligning with physician studies demonstrating significant reductions in note-taking time and documentation efficiency.^10,11,34^ Nearly all pharmacists interviewed felt time savings from ADT were underestimated by Epic Signal, which tracks the aggregate time of user activity for “Note and Letter” activities and navigators adjacent to documentation (i.e., reviewing labs, notes, and other information). Additionally, the activity timer starts/restarts for clicks, typing, scrolling, or other Epic interaction and pauses after 5 seconds of inactivity.^35^

### Pharmacist Perceptions of the Care Experience

‘Documentation Burden’ showed the greatest pre-/post-ADT improvement among all variables (Cohen’s d: -1.93). The findings were consistent with literature showing reduced documentation burden among physicians, despite generating longer and more detailed notes.^10,11,30,34,36^ This study’s qualitative data provided explanatory interpretation from pharmacists that reviewing and editing ADT-drafted notes required less T&E than writing their own, especially when documentation could not be started or completed during or immediately after encounters. Additionally, pharmacist interviews suggested the burden also shifted towards proofreading for errors (e.g., typos, inaccuracies), inclusion of only relevant information, and writing style preferences that parallel findings in the medical literature.^10,11^ Taken together, both data types indicated ADT substantially reduced document burden and reaffirmed the need for active pharmacist supervision (i.e., human-in-the-loop).

‘Patient Access’ showed no post-ADT improvement. The finding is consistent with literature reporting T&E savings from ADT do not translate to larger physician panels or more appointments per day.^26,37^ Pharmacist excerpts about whether they could add another patient to their schedule if urgently needed ranged widely. Taken together, the findings suggest that the large magnitude post-ADT improvements observed in the other care experience variables were either too small or unrelated to improve pharmacist perceptions of ‘Patient Access.’ The overall results also indicate that pharmacists face sizeable and persistent challenges related to time pressures, patient panel sizes, and administrative responsibilities like other healthcare professions.^38,39^

‘Undivided Attention’ improved post-ADT deployment (mean Δ: +25%), aligning with studies suggesting that physician T&E savings from diminished post-ADT documentation burden associated with better patient-centered care.^10,36^ In interviews, several pharmacists attributed improvements in other care experience aspects to increased patient connection, communication, and care quality. Even pharmacists reporting minimal or no self-perceived changes in their ability to give patients undivided attention, shared instances where ADT facilitated EHR interactions that improved quality care. The totality of the survey and interview data for this variable may suggest that more than producing a care experience that facilitates undivided attention, ADT enables pharmacists to practice in the way they see fit, or even prefer.

‘Afterhours Documentation’ generally improved for study pharmacists (mean Δ: -50%). The finding aligned with medical studies reporting post-ADT reductions in average afterhours work time, although results varied by physician characteristics or non-Abridge ADTs.^7,11,26,30,36^ Study pharmacist excerpts credited ADT for making work more manageable within regular clinic hours and a better quality of life.

The odds of pharmacists reporting any degree of ‘Burnout’ (Mini-Z ≥3) was 56% less likely post-ADT, but the finding was non-statistically significant. Some physician ADT-utilization studies failed to detected burnout improvements despite clinician-reported well-being improvements,^30^ but more evidence suggests that ADT significantly lowers burnout risk.^8,36^ Multiple studies indicate that greater levels of documentation burden and afterhours work positively relates with burnout,^40–42^ and most practice interventions take more than 6-12 months to detect burnout reductions on the Mini-Z instrument.^22,43–45^ Given several pharmacists expressed ADT improved quality of life aspects like less stressful workdays and work-from-home, more time is likely needed to better interpret this care experience aspect.

### Limitations

There are several limitations that should be considered when interpreting study findings. Threats to generalizability included being conducted a single health system and ADT licenses available to iOS users. Results also represent a narrow snapshot of one version of one product among several ADTs available on the market, in a rapidly evolving technological area. Thus, we adopted pre-existing measures from the ADT literature to improve comparability with past and future studies.

Another threat to external validity was oversampling of early adopters and individuals with strong ADT options due to a noncontrolled design. Thus, a purposive sampling strategy for diversifying pharmacist representation by practice subspecialty, time spent writing notes in the EHR, and years of MTDM practice to recruit pharmacists for Waves 1-3.

Threats to internal validity included known biases of pre-/post- surveys using self-report items (e.g., recall bias, social desirability bias, priming effects, attrition), EHR Signal logic (e.g., underestimation of total documentation time, lack of encounter level data), and semi-structured interviews (e.g., interviewer bias, inconsistent questioning). Thus, data source and method triangulation were used to enhance the credibility, comprehensiveness, and reliability of findings.

### Future Research

Future research with larger samples across multiple health systems and longer time horizons is needed to better understand ADT’s full and sustained impact on the pharmacist care experience. Future studies should also include patient-reported data and do head-to-head comparisons with human scribes or competing ADT products. Other potential variables for ADT studies are note quality, patient-clinician relationship impact, care continuity/coordination, and user or subspecialty phenotypes.

## CONCLUSION

In this mixed methods study, an ADT was associated with pharmacists spending less time in the EHR and perceptions of reduced documentation burden, greater ability to give patients undivided attention, and less afterhours documentation. Our findings suggest that ADT represents a promising technology for improving the pharmacist care experience.

## Disclosure of Conflict of Interests

The authors have no financial or other interests that could be perceived to bias their work. There are no financial support or personal connection with potential sponsors to disclose. Author SKD was both a participant and co-author for this study but did not have access to the data collected nor take part in the analysis.

## CRediT Statement

Olson: Conceptualization, Investigation, Methodology, Writing – Original Draft, Writing – Review & Editing, Supervision. Pradhan: Methodology, Validation, Formal Analysis, Investigation, Writing – Original Draft, Writing - Review & Editing. Whary: Investigation, Formal Analysis, Writing – Review & Editing, Project Administration. Dombrowski: Conceptualization, Participant, Writing – Review & Editing. Dobbins: Software, Writing – Original Draft, Writing - Review & Editing. Ward: Resources, Writing – Review & Editing, Project Administration. Obert: Investigation, Software, Resources, Writing – Review & Editing. Siegrist: Investigation, Software, Resources, Writing – Review & Editing. Wright: Conceptualization, Writing – Review & Editing, Supervision.

## Data Availability Statement

The data underlying this article cannot be shared publicly due to the privacy of individuals that participated in the study.

## Acknowledgements

The authors thank Salley Whitman and Jasmine Wong (Abridge) as well as Susanne Burns, Stacey Grassi, Jerry Greskovic, Josh King, Dan Longyhore, Amanda Popko, Josh Shaylor, Rebecca Stametz, David Vawdrey, Leeann Webster (Geisinger) for their technical expertise, data acquisition, and general advisements during the study. OpenEvidence was consulted to augment the connection between our findings and the literature.

## Notes

### Funding Statement

This study did not receive any funding.

### Author Declarations

The Geisinger IRB determined that this study met the criteria for a human subjects research exemption (Study#2025-0556) and gave ethical approval for this work.

